# Exploring the acceptability and adherence of the integrated MAMI care pathway for supporting vulnerable infants and their mothers in South Sudan: a qualitative study

**DOI:** 10.1101/2024.11.25.24317840

**Authors:** Hedwig Deconinck, Marlene Traore Hebie, Angelina Nasira Boi, Kirk Dearden

**Author notes:** **Corresponding author:** Kirk Dearden. Hedwig Deconinck, MAMI Consultant MOMENTUM Integrated Health Resilience, Institute of Health and Society, UCLouvain, Belgium Marlene Traore Hebie, Senior Technical Advisor, Nutrition, MOMENTUM Integrated Health Resilience, USA Angelina Nasira Boi, MAMI Coordinator, MOMENTUM Integrated Health Resilience, South Sudan Kirk Dearden, Technical Lead, Nutrition and WASH, MOMENTUM Integrated Health Resilience, USA.

## Abstract

**Background:** One in four infants is born too small or too early, increasing the risk of poor growth, development, ill health, and death. This risk is exacerbated by recurring conflict and climatic shocks. The management of small and nutritionally at-risk infants under 6 months and their mothers (MAMI) care pathway was integrated into primary healthcare to ensure continuous mother-infant-centered care in South Sudan. This study explored the acceptability of the MAMI approach for service providers and users and the factors influencing their adherence.

**Methods:** A qualitative study was embedded in a mixed-methods study following 521 vulnerable mother-infant pairs. MAMI care included sensitization and screening in the community and health facility, in-depth assessment, and risk-based care addressing physical, mental and nutritional vulnerability of infants and mothers. Semi-structured interviews of 20 health workers and 30 mothers explored acceptability and adherence to MAMI care provision and use.

**Results:** Themes of perceived acceptability of MAMI care were mapped across coherence, self-efficacy, affective attitude, burden, ethicality, opportunity cost, and perceived effectiveness. Health workers were most apprehensive about the additional workload and time required to implement the care pathway. Mothers expressed concern over the numerous health visits far from their homes and not having supportive family members. Factors promoting adherence to MAMI care provision and use were mapped across capability, opportunity, and motivation. Health workers’ self-confidence was boosted by the new knowledge and skills they acquired through training and mentorship. Mothers reported learning practical ways to care for their infants independently and to witness the positive impact on their infants’ thriving.

**Conclusion:** MAMI care was accepted for addressing the needs of a previously overlooked vulnerable group. To ensure effective adoption, delivery and use, challenges must be resolved. Adapting the MAMI care pathway to the local health system context will encourage adherence from both providers and users.

**Key messages:**

- **What is already known on this topic** – The 2023 WHO updated guideline on preventing and treating wasting in children under 5 includes care for infants under 6 months at risk of poor growth and development and their mothers within primary healthcare. Understanding and effectively delivering care to this previously overlooked vulnerable group is critical for national policymakers and implementers to address persistently high levels of infant mortality.
- **What this study adds** – This study explored the acceptability of the MAMI care pathway–a risk-based, mother-infant centered care approach–among service providers and users in South Sudan, as well as the factors influencing their adherence. It offers recommendations for effectively adapting the approach to the local health system context to promote adherence from both providers and users.
- **How this study might affect research, practice or policy** – The study generated valuable insights and learning at both national and global levels to enhance policies and practices. It also laid the foundation for future research and intervention development, particularly in settings with similar demographics and healthcare challenges.

## Introduction

Globally, an estimated one in four infants under 6 months old (u6m) is small or nutritionally at risk, leading to poor growth and development, ill health and death (1, 2). Although deaths in children under 5 years old have declined in South Sudan, children with wasting have a nine times higher risk of death than healthy children, overburdening the health system (3).

National maternal, neonatal and child health and nutrition policies address vulnerability in infants u6m and their mothers using various approaches. Established primary care practices for infants u6m include vaccinations at birth and at 6, 10 and 14 weeks, with over 70 percent coverage against common childhood illnesses (6), and health seeking for ill infants. Maternal care and support are fragmented across health units, and there is a lack of psychosocial and mental health services, feeding and care counselling and early childhood development (ECD) (4). Moreover, irregular and inadequately covered technical and financial support hinders effective health service delivery (4, 5).

To address care challenges for vulnerable mother-infant pairs, Momentum Integrated Health Resilience (MIHR) partnered with the South Sudan Ministry of Health (MOH) in 2022 to adapt and field test the management of small and nutritionally at-risk infants u6m and their mothers (MAMI) integrated care pathway (7, 8). The MAMI care pathway (henceforth MAMI care) delivered in primary care consisted of risk screening in both communities and health facilities, in-depth assessment and risk classification and targeted risk-based care and counselling for physical and mental health and nutrition of both infants and mothers (12, 13). The exercise aimed to generate national and global learning to improve policies and practices.

During implementation of MAMI care, challenges and outcomes were noted by the study team and discussed but not always fully understood. This study explored the acceptability of MAMI care to both health workers and mothers to understand what worked for whom under what circumstances and to identify factors that facilitated or hindered adherence to care by service providers and users (9).

## Methods

### Qualitative approach and research paradigm

The theory-driven qualitative study used Sekhon’s theoretical framework of acceptability (11) and Michie’s Capability, Opportunity and Motivation Behavior Model (COM-B) (9) to explore acceptability and understand adherence behavior of key informants in semi-structured interviews.

### Context

The study was conducted in February–March 2024 in five sites in Central Equatoria, Jonglei, Western Bahr el-Ghazal and Western Equatoria states in South Sudan, nested in a longitudinal multi-methods study of 521 vulnerable mother-infant pairs enrolled and supported between October 2022 and December 2023 (15 months) in the MAMI CP until the infants reached 6 months of age. Each site had an MOH-run primary healthcare center (PHCC), a referral hospital and a County Health Department (CHD) for oversight.

### Researcher characteristics

Two international study investigators supported a local lead interviewer, an assistant and a boma (community) health worker (BHW) as translator with proficiency in English and local languages.

### Sampling strategy and units of study

Fifty key informants—20 MAMI care providers and 30 MAMI care users—participated in semi-structured interviews. Care providers included five MAMI assistants who were previously MIHR staff and supported one of the five MAMI implementation sites, five clinical officers who were MOH staff and integrated MAMI care as part of their regular consultation duties and 10 BHWs appointed by their communities and supported by MIHR to integrate MAMI care in the catchment areas of the MAMI sites included in the study. Gurei and Pariak sites were purposively selected to represent an urban and a rural context. The clinical officers were purposively selected based on their involvement in MAMI care. The BHWs were randomly selected from a list of eighteen MIHR-supported BHWs. Thirty mothers were randomly selected from the MAMI individual care database and stratified in three adherence categories: Group 1 attended all follow-up visits until exit, group 2 returned at least once for a follow-up visit, and group 3 discontinued care immediately after enrolment. All participants were informed of the purpose, date and duration of the interview and received money for transport to the interview site. The number of key informants was sufficient to obtain variation and reach saturation. All care providers had received initial training and ongoing mentoring on the MAMI care before implementation, and all mothers had agreed to enroll in the care pathway after providing informed consent.

### Ethical approval

This study is part of the overall MAMI implementation study protocol that received ethical approval from the Institutional Review Board (IRB) of Momentum-JSI’s and the Ministry of Health of South Sudan in March 2022.

### Data collection methods, instruments and technologies

Interviews were conducted face-to-face whenever possible, and due to travel issues, some were conducted online using Microsoft Teams. Both study investigators participated in all interviews online. In-person interviews with health workers and mothers took place at PHCC sites. Interviews followed semi-structured interview guides and lasted on average 90 minutes. Discussions were held in English, Arabic or the local language with simultaneous translation.

### Data processing and analysis

The study team debriefed after each interview on key points and clarified inconsistencies in information. Interviews and discussions were recorded, transcribed using otter.ai and summarized by theoretical constructs in Excel. Demographic and care information was retrieved from interviews and study databases. Recurrent themes were coded and analyzed from care providers’ and users’ perspectives. Two study investigators discussed coding consistency across the interview data. The acceptability framework included seven constructs: coherence, self-efficacy, affective attitude, burden, ethicality, opportunity cost and perceived effectiveness. Adherence behavior factors were mapped using the COM-B model’s capability, opportunity and motivation.

### Techniques to enhance trustworthiness

Data were triangulated and iterated for consistency of interpretation of discussions within and across themes, providing reliable and valid insights.

Research processes and decisions were recorded in detail for study transparency and reproducibility. Reflective journaling recorded biases, alterations and decisions made during the study to improve confirmability and identify and document recurring and deviant patterns. The structure of this manuscript follows the Standards for Reporting Qualitative Research (SRQR) checklist (10).

## Results

### Characteristics of the MAMI care providers and users

Five of the 20 participating health workers were MIHR-employed MAMI assistants, and five were MOH-employed clinical officers (two of the five were officers-in-charge of the PHCCs. Each MAMI assistant and clinical officer represented one of the five MIHR-supported MAMI implementation sites. All clinical officers involved in MAMI care had clinical nurse education and four were male. Ten community-appointed BHWs receiving incentives from MIHR were from urban Gurei and rural Pariak and six were male.

All 30 mothers were the biological mothers of the infants, and half resided in Gurei or Pariak. Only about one-third had received any education. More mothers in group 1(younger, less educated and more involved in farming), delivered in the maternity unit. Key vulnerability factors related to the mother for enrolment in MAMI care were primiparity, followed by lactation difficulties and mid-upper arm circumference (MUAC) below (<) 230 mm. These mothers completed on average three follow-up visits, stayed longer in care (24 weeks), received more support from their mothers (or mothers-in-law) or husbands and advised other mothers about MAMI. Maternal mental health measured with the Patient Health Questions-9 score was low across the three groups (score below (<)10). Although some mothers reported knowing the BHWs in their communities, few had received explicit support from them. The most common point of referral for MAMI was the vaccination unit. Most of the 13 mother-infant pairs who ended care when the infant reached 6 months of age recovered, i.e., not requiring follow-on healthcare services (Table 1).

**Table 1:** Characteristics of mothers participating in the South Sudan study, February– March 2024.

| Characteristics of mothers and infants of enrolled pairs | Group 1<br>Follow-up visits<br>until 6m of age | Group 2<br>Few follow-up<br>visits | Group 3<br>No follow-up visit | All* |
| --- | --- | --- | --- | --- |
| All | 13 | 8 | 9 | 30 |
| Urban (vs rural) | 8 | 2 | 5 | 15 |
| **MOTHER** |  |  |  |  |
| Mother (vs caregiver) | 13 | 8 | 9 | 30 |
| Mother's age at enrolment (years) (median) | 20.0 | 26.0 | 25.0 | 200 |
| Mother's education: |  |  |  |  |
| None | 10 | 6 | 6 | 22 |
| Primary | 2 | 1 | 2 | 5 |
| Secondary | 1 | 1 | 1 | 3 |
| Mothers' source of income: |  |  |  |  |
| Commerce | 2 | 2 | 1 | 5 |
| Farming | 7 | 2 | 5 | 14 |
| Housekeeping | 4 | 4 | 3 | 11 |
| Mothers' access to health services: |  |  |  |  |
| Attended ANC | 4 | 1 | 5 | 10 |
| Delivered at maternity unit | 5 | 2 | 3 | 10 |
| Mother's MUAC at enrolment (mm) (median) | 256 | 238 | 250 | 256 |
| Mothers' reasons for being classified as at moderate risk: |  |  |  |  |
| Low MUAC (<230 mm) | 3 | 1 | 1 | 5 |
| Lactation difficulties | 3 | 2 | 2 | 7 |
| Primipara | 9 | 3 | 4 | 16 |
| Multipara | 1 | 2 | 1 | 4 |
| Adolescent mothers | 2 | 1 | 1 | 4 |
| Maternal mental health (N=25): |  |  |  |  |
| Mental health low risk (PHQ-9 score <10) | 10 | 7 | 8 | 25 |
| Mental health moderate or high risk (PHQ-9 score ≥10 or positive Question9) | 0 | 0 | 0 | 0 |
| Mothers in MAMI care shared experiences with others: |  |  |  |  |
| Mothers advised mothers (one to one) | 9 | 4 | 4 | 17 |
| Mothers shared MAMI information in women's groups | 3 | 0 | 0 | 3 |
| Mothers receiving support: |  |  |  |  |
| Support from mother (or in-law) | 7 | 2 | 2 | 11 |
| Support from family | 1 | 1 | 4 | 6 |
| Boma health worker (BHW) known | 4 | 1 | 4 | 9 |
| Support from BHW | 1 | 0 | 1 | 2 |
| Mothers in MAMI care shared experiences with others: |  |  |  |  |
| Mothers advised mothers (one to one) | 9 | 4 | 4 | 17 |
| Mothers shared MAMI information in women's groups | 3 | 0 | 0 | 3 |
| Number of return visits after enrolment (median) | 3 | 2 | 0 | 3 |
| Duration of care (enrolment to last visit) (weeks) (median) | 24 | 14 | 0 | 15 |
| Care outcome: |  |  |  |  |
| Recovered | 11 | NA | NA | 11 |
| Not-recovered | 2 | NA | NA | 2 |
| **INFANT** |  |  |  |  |
| Male infants | 5 | 4 | 5 | 14 |
| Infant age at enrolment (months) (median) | 1.7 | 0.4 | 1.0 | 1.4 |
| Infant's MUAC at enrolment (mm) (median) | 108 | 100 | 107 | 104 |
| Infants' reasons for being classified as at moderate risk: |  |  |  |  |
| Mother-reported low birthweight (LBW <2500 g) | 0 | 1 | 0 | 1 |
| Mother-reported preterm birth (<37 weeks gestation) | 0 | 0 | 1 | 1 |
| Morbidity | 7 | 5 | 5 | 17 |
| Low MUAC (Infant <7 weeks MUAC <110 mm) | 5 | 5 | 5 | 15 |
| Low MUAC (Infant ≥7 weeks MUAC <115 mm) | 4 | 3 | 2 | 9 |
| Low WAZ (<-2) | 0 | 3 | 0 | 3 |
| Feeding problem | 13 | 11 | 6 | 30 |

\* Note that for some indicators the sample size is smaller than 30 due to missing data.
ANC= antenatal care; BHW= boma health worker; MUAC= Mid-upper arm circumference; NA= not applicable; PHQ-9= mental health assessment tool.

Infants in group 1 were older on average on enrolment. Across the three groups, almost all infants <7 weeks had a MUAC <110 mm, and two-thirds of infants above or equal to (≥) 7 weeks had a MUAC <115 mm. Key vulnerability factors related to the infant for enrollment were feeding problems, morbidity and low MUAC (Table 1).

### Thematic analysis of acceptability

Analysis of the perception of MAMI care among health workers and mothers yielded 29 and 33 themes, respectively, across the seven theoretical constructs of healthcare acceptability, some of which were repeated when a different construct lens was applied (Figures 1 and 2).

**Figure 1:**
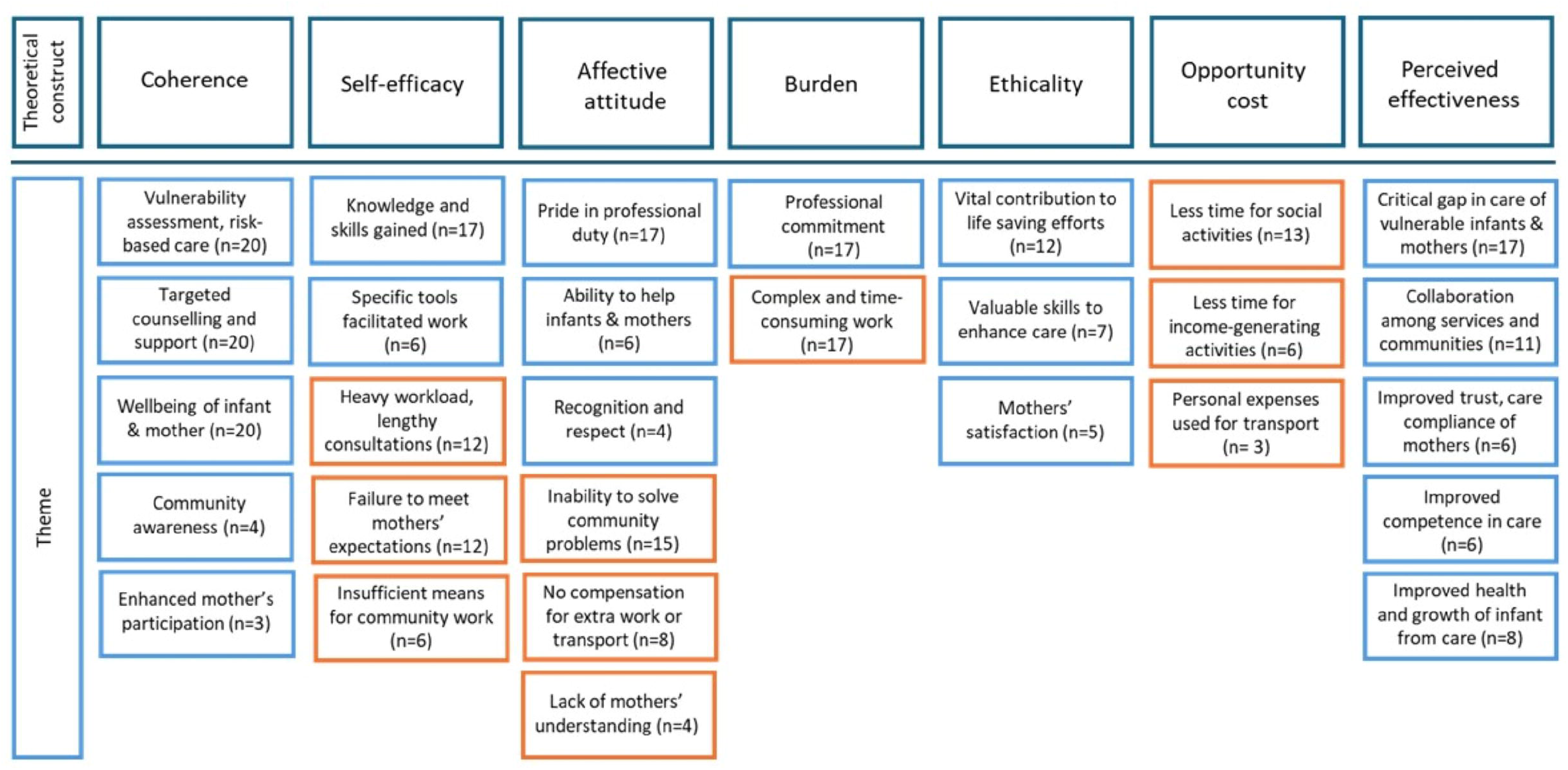
Themes of acceptability of the MAMI Care Pathway approach as perceived by care providers participating in the South Sudan study, February–March 2024 (blue boxes are positive or enabling factors, and orange boxes are negative or hindering factors).

**Figure 2:**
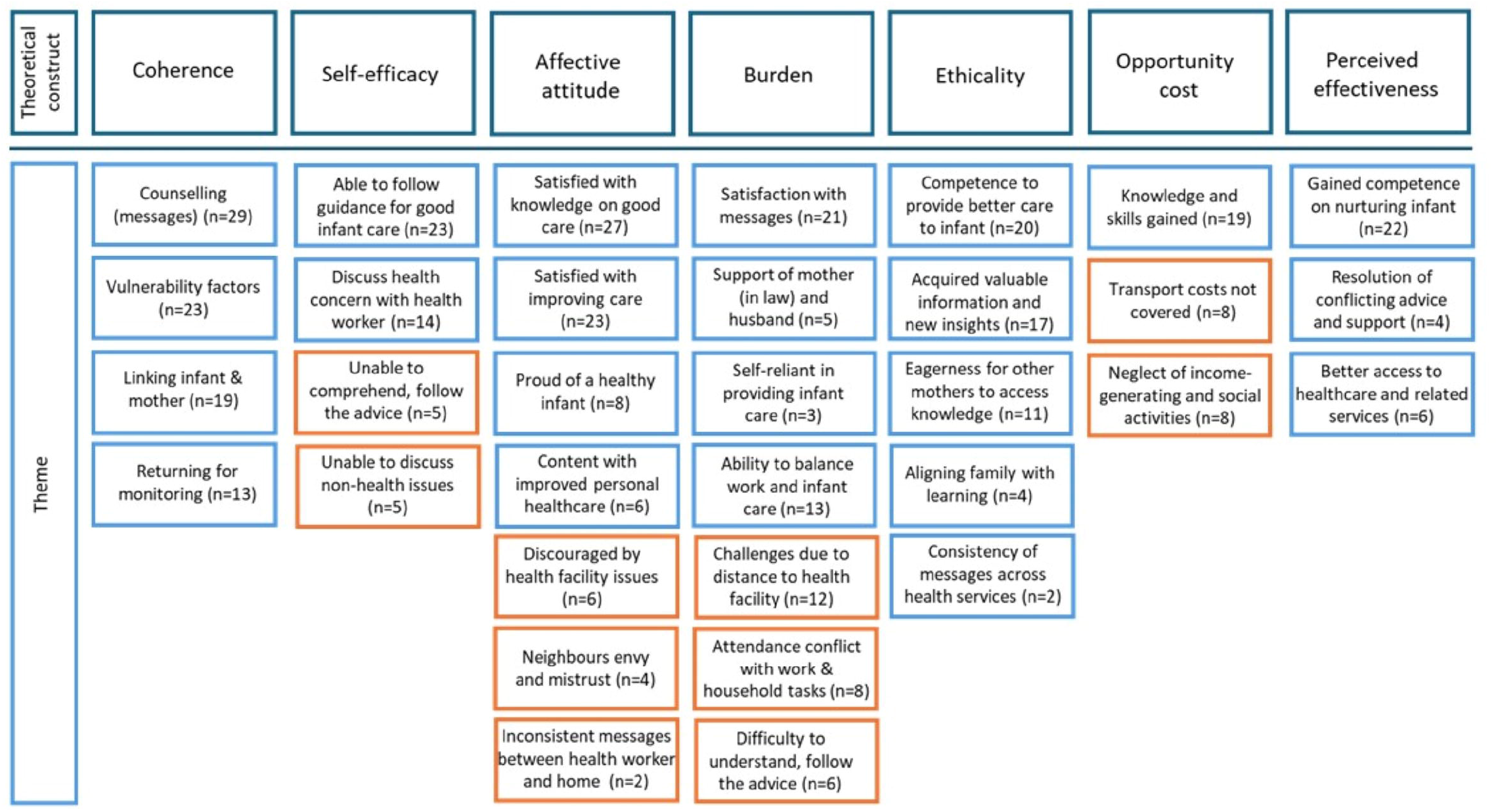
Themes of acceptability of the MAMI Care Pathway approach as perceived by care users participating in the South Sudan study, February–March 2024 (blue boxes are positive or enabling factors and orange boxes are negative or hindering factors).

### Health workers’ perceptions of care

#### Coherence, or understanding MAMI care and how it worked

The coherence construct focused on how health workers described MAMI care and which aspects they found important or new. All health workers described MAMI care as encompassing vulnerability assessment and risk-based care for infants u6m and their mothers through targeted counselling and support. They noted that since the introduction of MAMI care, the vulnerability of small, sick newborns and infants under 6 months has been acknowledged. Previously, a breastfed newborn was considered stable and only needed to return to the PHCC for vaccination or when ill.

> *“Before, when delivering a stable newborn, it was assumed that everything was OK, and they would only return after 6 weeks for EPI … but now with MAMI, following growth and development closely, this was an eye opener.” MAMI Assistant 3*

Some health workers mentioned the importance of community awareness to encourage mothers.

> *“Sensitization in the community is important in supporting mothers in knowing and receiving family support to be able to go to the HF and follow the advice.” MAMI Assistant 2*

#### Self-efficacy, or confidence in providing MAMI care

Most health workers felt competent to assess vulnerability and give mothers targeted messages about healthy behaviors.

> *“I was able because the training was perfect, and MAMI became easy, I was able to do in my daily activity. Previous knowledge plus new knowledge helped me*…*” Clinical Officer 5*

Some liked the tools that facilitated their work.

> *“Assessing using the form is better because the questions are there, and the findings are recorded and cases are identified. If referral, … without forms, this information is written in the mother’s ‘book. The form is far better and fast than being writing on any paper.” Clinical Officer 4*

On the other hand, health workers complained about the heavy workload, lengthy consultations and lack of teamwork and links with the hospital to follow up referred mother-infant pairs.

> *“Whatever you do many times comes easy. IMCI assessment was easy because I have been doing it before. What was challenging was time and workload and working alone.” Clinical Officer 2*

Several felt the lack of transport to the communities or incentives hindered their ability to convince mothers to be involved in MAMI care.

> *“Mothers have a lot of expectations and challenges to address. They expect drugs or other tangible support, and we have nothing to give.” MAMI Assistant 4*

#### Affective attitude, or interest in being involved in MAMI care

Most health workers felt good about providing care to a previously ignored but vulnerable population. Some felt they could facilitate mothers’ access to care across services and in turn receive recognition and respect.

> *“I was touched by being involved with MAMI because no one talked about this very vulnerable population, which was left out, and we provided services and linked with other services. … Mothers were touched by the messages. For example, expressing milk for mothers when they had to be absent, was unheard of. They would rather give cow milk.” MAMI Assistant 2*

Others felt frustrated when they failed to solve mothers’ problems.

> *“Mothers were comfortable to share health-related issues. It is good to talk, and if they understand they will adhere and if not, they will not. There are a lot of misconceptions, cultural taboos and certain topics are difficult to provide. … Also in our tradition it is not good to enter a house where a mother has delivered, sometimes I feel uncomfortable to assess a newborn, unless you give something good.” Pariak BHW 2*

Nearly half of the health workers were disappointed in the lack of compensation for the extra work and transport to provide MAMI care.

> *“Discouraging is that I work but am not paid.” Pariak BHW 4*

#### Burden, or efforts to provide MAMI care

Some health workers did not find MAMI care a burden because they saw it as their professional commitment and were motivated by what they learned and the technical support they received during implementation.

> *“The collaboration and learning from each other was good. Collaboration with the MAMI Assistant helped to start early with the MAMI cases … The risk assessment needs a good mind set.” Clinical Officer 5*

On the other hand, they felt MAMI care was complex and time-consuming and felt they should be paid an incentive for the extra work. Although they could see the purpose of the forms and paperwork, many found them a burden.

> *“The forms were too long, and it takes longer hours and many mothers on the line were complaining. I feel like I’m not doing enough and faster while conducting risk assessment.” Clinical Officer 5*

#### Ethicality, or whether MAMI care fits with the value system

Most health workers recognized that MAMI care saved lives and found fulfilment in aiding a vulnerable but previously neglected population. Acquiring skills to serve vulnerable mother-infant pairs bolstered their confidence and engagement. Positive feedback from mothers reinforced their belief in the value of their work.

> *“Meaningful good messages that prevent death. What is different now is that MAMI also focuses on u6m.” Gurei BHW 3*

#### Opportunity cost or sacrifices and gains from providing MAMI care

Most health workers regretted having less personal time or time for income-generating activities because of longer working hours.

> *“Before, I was a businesswoman in the market. Since I became a BHW, I stopped. … I work more than four hours and have to give up income and social contact. I’m not able to cultivate. I cannot go and meet my friends. When I reach home, it is late.” Gurei BHW 2*

#### Perceived effectiveness, or the ability of MAMI care to achieve its purpose as expected

Most health workers felt MAMI care addressed a vital need by offering specialized, risk-based care across services and in the communities. About one-third appreciated that MAMI care enhanced infant health and development, which in turn increased maternal trust and adherence to care practices. This improvement in care adherence had tangible results.

> *“MAMI filled a gap. Nobody paid attention to u6m before… For the infant, there was vaccination and concern in case of illness. MAMI makes an interconnection … a collaboration between maternity, EPI, OPD and the movements between them.” MAMI Assistant 2*

### Mothers’ perceptions of care

#### Coherence, or understanding MAMI care and how it worked

The coherence construct focused on what mothers remembered about the MAMI Care Pathway and which messages they found important. Most mothers understood that MAMI care was about identifying vulnerability factors, receiving targeted counselling to improve infant health and growth and taking good care of themselves. Messages they retained included exclusive and frequent breastfeeding up to 6 months, maintaining good hygiene, and returning to the health facility when their infants were sick. For the mothers, counselling meant “*listening to and following up health messages”*.

> *“MAMI is assessing infant and mother to know their health, for taking care … In MAMI there are pictures of breastfeeding and position, and on hygiene… My family told to give water to the infant, water is good when the infant is thirsty. In MAMI I got the message to do exclusive breastfeeding, because breastfeeding includes water.” Mother GUR08 Group*

#### ‘Self-efficacy’, or confidence in applying the advice received in MAMI care

Most mothers were grateful for the advice because they wanted to take good care of their infants and managed to follow the advice. Receiving the same messages across healthcare units reinforced the messages.

> *“I was confident to follow the [new] advice of the health worker, and even when my own mother had other ideas, but when I informed her, she accepted.” Mother PAR05 Group 1*

Most mothers were unconcerned about the gender of the health workers. However, they did not consider psychological stress or mental wellbeing as health issues, and therefore did not discuss them with health workers. Family or couple conflicts were kept within the family.

> *“Personal issues I cannot discuss … because I feel ashamed to share because these are family issues, not health issues, so I cannot share.” Mother PAR05 Group 1*

A few mothers did not follow the advice because they misunderstood MAMI care, were not told clearly what it was about or were not motivated. One deliberately ignored the advice to attend supplementary feeding because she did not like the food given. Some mothers felt they were wasting their time when they did not receive any tangible incentive in MAMI care.

> *“ … I expected to be given milk… I am selling tea. I leave my infant at home with infant formula. My job does not allow me to take the baby*… *I was told to return after one month. I did not return. I was wasting my time.” Mother GUR09 Group 3*

#### Affective attitude, or interest in being involved in MAMI care

Mothers were grateful for the messages they received and appreciated the skills they acquired to better care for their infants. Even mothers who never returned or returned only a few times appreciated the messages, listened and applied the advice.

> *“I got good messages when I needed it most, this made me very happy. … Messages on “Take good care of baby, take good care of yourself” helped me a lot.” Mother GUR37 Group 1*

Many mothers mentioned that the messages were new to them, and some enthusiastically recalled their MUAC classification.

> *“My child was OK but I was thin, my size was less than 230.” Mother PAR74 Group 3*

The novelty of attention to the mothers and the explicit link between infants and mothers made the mothers feel supported.

> *“I always feel good [about MAMI care], … it gave important information that I did not know. … I liked that I was given messages which were very crucial which can really help a mother on how to take care of the baby and you yourself as a mother.” PAR19 Group 2*

MAMI care was especially appreciated by first-time mothers, who usually felt unskilled and were dominated by older women.

> *“My positive feeling is that I feel happy about MAMI because I got good messages and when I put them into practice result in having good health of the baby, so I felt happy about it.” Mother PAR62 Group 3*

Less than half of the mothers felt returning for follow-up visits to monitor their infants’ growth and their own wellbeing important. Some said they did not return because they were not told. But most mothers who went to the PHCC did so for their infants’ vaccination, a well-established healthcare service mothers adhered to.

> *“My family members were supportive to come for vaccination. When it is time for me to come for vaccination, they can give me transport fee to use motorcycle to come.” Mother PAR74 Group 3*

Some mothers found the number of questions and unsolicited attention they received when they came to the PHCC for vaccination worrying and confusing.

> *“I was not used to all these questions and I felt worried, as if there was something wrong with me and my baby. … I saw MUAC taken of older kids, but not so small as mine.” Mother GUR51 Group 2*

Some mothers were unhappy about long lines at the PHCC, drug stockouts that meant having to buy medicines from local vendors, lack of incentives such as soap or mosquito nets and the need to pay for notebooks to keep consultation notes as a condition of access to the PHCCs.

> *“I like the care and receiving drugs. I do not like the long waiting time, and there is no water to drink… If you do not pay for the book, then you cannot see the doctor.” Mother GUR 18 Group 3*

#### Burden, or efforts to adhere to MAMI care and advice

Most mothers felt that the advice to support their infants’ health made their participation in MAMI care less burdensome. Some were relieved by encouragements from mothers (mothers-in-law) or husbands.

> *“Not a burden to listen to messages and follow the advice. I believe that has helped me, changed my life, now I know how to take care in the right way.” Mother GUR70 Group 2*

Concerns were mostly related to distance to the PHCC, time and travel costs, having to leave other children at home, missing work or household duties, being criticized by household members or inquisitive neighbors and insecurity of travel in some areas. Some mothers did not understand the advice or could not follow it.

> *“I have no transport, I came on foot, I am at home alone with the children, so I cannot come as I would like.” Mother GUR94 Group 2*

#### Ethicality, or whether MAMI care fits with the value system

Most mothers felt able to provide better care for their infants, the most valuable perceived benefit of MAMI care. Their gratitude for new knowledge and skills made them want other mothers to benefit. A few said that consistent messages across healthcare units made them feel secure and confident.

> *“… I would not have known …, the women in my home would not have told me … I could not have taken care of myself. I delivered in the PHCC, and I received the same messages there when I came for antenatal care and delivery.” Mother PAR49 Group 2*

#### Opportunity cost, or sacrifices and gains from adhering to MAMI care

Mothers felt they gained knowledge and skills from MAMI that they would never have learned or obtained elsewhere.

> *“It was possible to return [four times]. I was motivated to come, each time I received a check and encouragements. Even neighbors started asking: “where do you go and why”?*

> *And answered, “To improve the health of my baby”. On each visit, it was great to have direct access to the doctor”. Mother GUR73 Group 1*

Conversely, regardless of their adherence to follow-up visits, many mothers faced difficulties securing transport or had to forgo income-generating and social activities.

> *“Visits to the PHCC are difficult because it is far, and I have to leave things at home. … MAMI messages were helping me as a mother … but MAMI consumed too much of my time… I had to give up important things such as farming, going to watch wrestling, meeting friends.” Mother PAR19 Group 3*

#### Perceived effectiveness, or the ability of MAMI care to achieve its purpose as expected

Mothers strongly affirmed that the skills they gained made their infants healthier.

> *“MAMI helped me and my child and we are living better. My child is healthy not being sick no diarrhea. I see child of my brother with lot of diarrhea.” Mother PAR09 Group 1*

A small proportion felt that participation in MAMI care facilitated access to health workers, who were more attentive to their concerns. Others were pleased to resolve conflicting advice from the PHCCs and their families.

> *“Some neighbors advised me to go and see a traditional healer, but I said no. On each visit, it was great to have direct access to the doctor…Now my daughter is healthy, which I could not have done with advice from neighbors.” Mother GUR73 Group 3*

### Thematic analysis of adherence behavior

Factors that influenced adherence to MAMI care were mapped on the COM-B framework of Michie et al. (Table 2) (9). The framework proposes three conditions that interact to generate behavior that in turn influences these conditions. Capability is knowledge and skills to engage in an activity, both psychological (ability to engage in thought processes, capacity to plan) and physical (skill to undertake and follow advice). Opportunity is factors that enable or prompt the desired behavior, both physical (afforded by the environment) and social (afforded by the cultural context, supportive network). Motivation is attitudes and aspirations that energize and direct behavior, both automatic (involving emotions and impulses) and reflective (involving evaluations and plans) (9, 14).

**Table 2:**
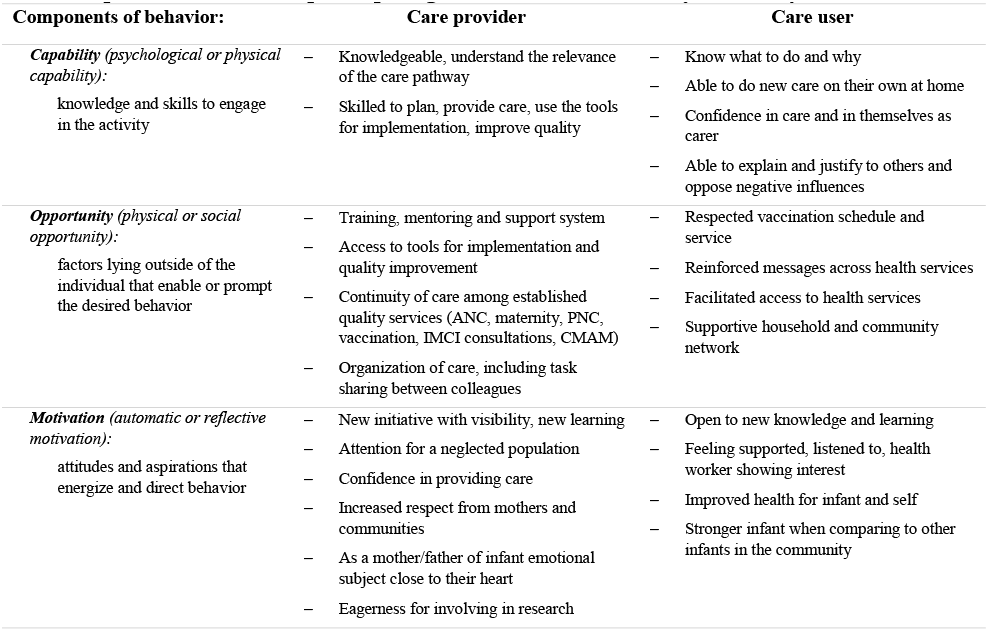
Map of influencing factors of positive adherence behavior as perceived by MAMI care providers and users participating in the South Sudan study, February–March 2024.

The findings of the thematic analysis in the COM-B framework show drivers of adherence to MAMI care (Figure 3). For health workers, the key factor in adhering to care was self-confidence and the ability to care for a previously neglected vulnerable population. For mothers, the key factor was seeing their infants thrive.

**Figure 3:**
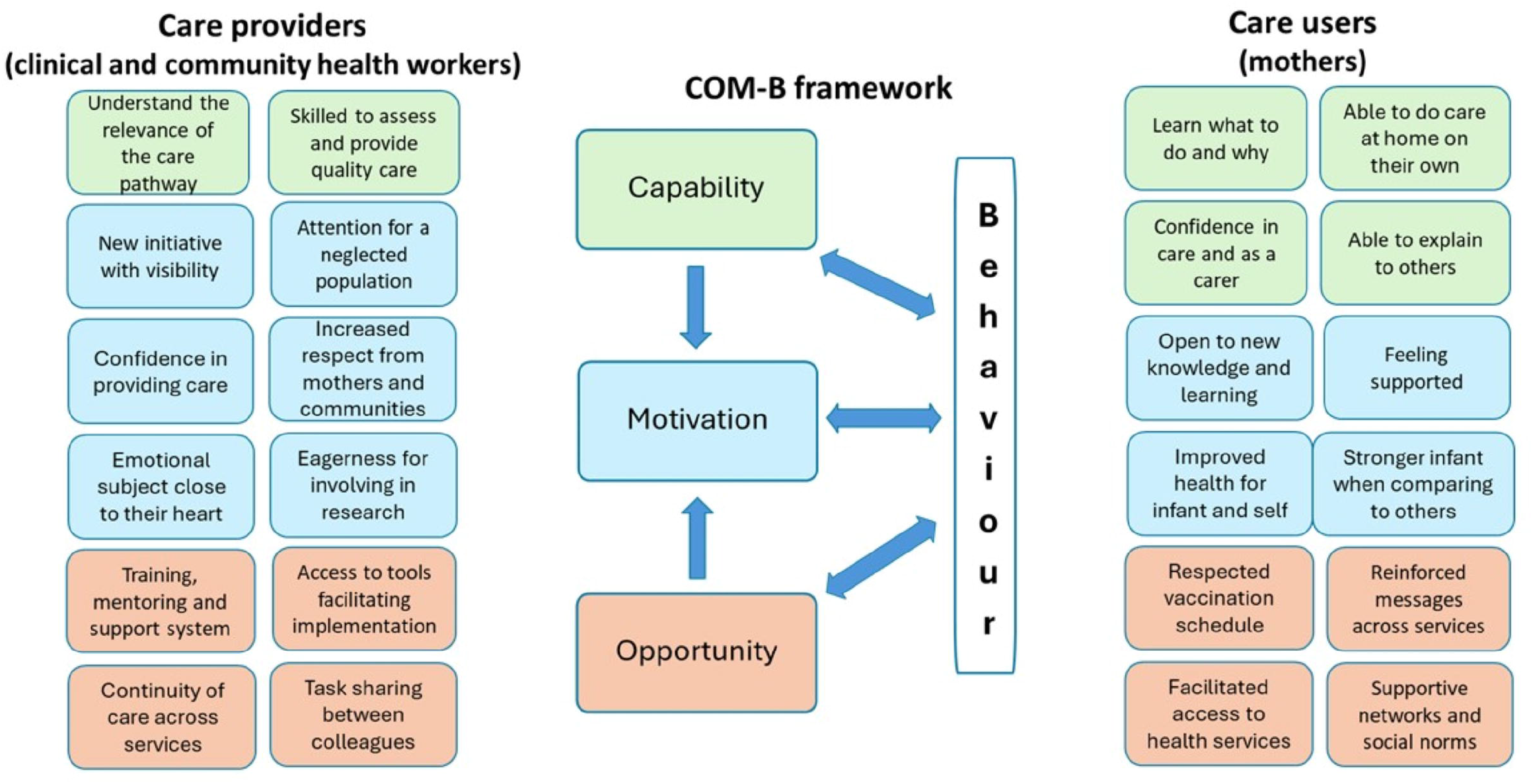
Behavior conditions that improve adherence to MAMI care provision and use as perceived by MAMI care providers and users participating in the South Sudan study, February– March 2024.

## Discussion

The study enhanced understanding of the effectiveness of MAMI care by examining its acceptability among health workers and mothers of enrolled mother-infant pairs and identified factors that facilitated or hindered adherence to MAMI care.

Health workers were trained and shaped by vertical health programs that lacked a holistic approach, resulting in gaps in continuity of care. For example, they often overlooked infants u6m after a healthy start, assuming that breastfeeding would proceed smoothly, common diseases would be infrequent, and growth and development issues would only arise with the introduction of complementary feeding and exposure to unhealthy environments.

Gaining the expertise and skills to serve this vulnerable group enhanced health worker’s sense of professional fulfilment. However, they did not report on challenges in addressing maternal physical and mental health, an aspect the interview did not uncover. Cultural norms do not recognize psychological and mental problems as health issues or allow open discussion.

Clinical health workers worried most about the additional workload for in-depth assessment, filling out forms and providing risk-based counselling. The issue, however, seemed to be lack of routine assessment. With good practice, this activity would be fast, as few mother-infant pairs would present risks requiring intensive attention. Also, effective organizational skills could help manage crowds more efficiently, distribute consultations more evenly or extend the activity window beyond 9 am to 12 pm. In some sites, MAMI screening and assessment slowed when the MAMI assistants or the one trained clinical officer, who were exclusively assigned these tasks, were absent. Vulnerability screening focused heavily on measuring MUAC, which was easy, while key vulnerability questions received less attention. This aligns with mothers’ reports that community nutrition volunteers only measured MUAC of children 6–59 months, overlooking other important health and nutrition issues.

BHWs trained and equipped to implement community-based healthcare, which included MIHR-specific maternal and child health activities like MAMI, had mixed feelings about MAMI care. The extra workload was difficult to fit in their voluntary work schedule, and cultural norms made access to households with newborns difficult. Nonetheless, the BHI materials covered most vulnerabilities for infants u6m and their mothers, except for measuring MUAC in infants u6m and the mother-infant pair-centered care approach. BHWs were often too busy or did not focus on infants u6m, as the BHI targets children under 5 years for vaccination, illness or malnutrition for 6–59 months. Newborns are kept inside the house, where BHWs are not allowed without the father’s permission. Consequently, small infants were not their primary concern.

Mothers were eager to learn how to care for their infants and pleased when their infants thrived when they followed MAMI advice. However, targeted counseling was often simple message sharing, which might not be enough to change behavior. Mothers adhered to advice if it seemed feasible and had family support, facilitated by community awareness. Urban mothers reported less family support and social interaction than rural mothers. Mothers had difficulty discussing mental health issues, which were much misunderstood. All mothers were concerned about frequent follow-up visits, especially traveling to PHCCs far away with newborns or small infants. Transportation costs, mentioned as a major barrier to follow-up visits, often masked more complex reasons. Mothers sometimes felt the trip was not worth the effort compared to other NGO-supported programs that provided incentives. However, adherence to the vaccination schedule was well respected and supported by the community, suggesting that MAMI follow-up schedules could align with these appointments to improve adherence. Also, the contact with the BHW seemed limited to measuring MUAC of older children, leaving young mothers in a vacuum in terms of infant feeding and care. It is no surprise mothers were eager for MAMI care to continue.

Missed opportunities for health workers included adequate time and skills for counseling, addressing maternal mental health, community engagement, consultation management, form proficiency, and task sharing. For mothers, it included connections to BHWs and adequate family and community support. Seized opportunities were the strengthened integrated management of neonatal and childhood illness (IMNCI) approach and engaging mothers in their infants’ health. Recommendations for improvement included making MAMI care routinely available, improving patient flow, combining visits, and giving consistent health advice. Follow-up visits should be mutually agreed upon with mothers, considering their ability to return for care. In the community. BHW teams should adopt a holistic, person-centered approach, decentralize health care, and strengthen individual counseling. Urban mothers would benefit from health education through radio, theatre, telephone, and television.

This study faced limitations. First, the interviews were conducted by the MAMI coordinator, who was also an employee of MIHR and responsible for overseeing MAMI implementation. This overlap in roles could have affected participants’ willingness to speak freely, as well as the coordinator’s impartiality during the interviews (interviewer bias). Second, interviews were mainly done at the MIHR office or the PHCC. Health workers, still under MIHR employment, might have been less willing to be frank in their responses. Mothers interviewed at their PHCCs might have felt pressured to provide responses they perceived as socially acceptable or favorable. Third, interviews with mothers were conducted in their native language and immediately translated by a proficient boma health worker. Despite the translators’ expertise, subtle nuances might have been lost, necessitating careful interpretation of quotes (translation accuracy). The interview team and two study investigators addressed these limitations during interview debriefings to minimize the impact of biases.

## Conclusion

The study found the MAMI Care Pathway acceptable to both providers and users, providing care for a previously overlooked vulnerable group. It highlighted the need for policy changes to prioritize comprehensive, patient-centered care for vulnerable infants u6m and their mothers. To ensure effective adoption, delivery and use challenges must be resolved. Successful adaptation will encourage adherence from both providers and users.

The findings, though context-specific, may be applicable to other settings with similar demographics and healthcare challenges, and provide a foundation for future research and intervention development.

## Data Availability

All data produced in the present study are available upon reasonable request to the authors.

## Acknowledgements

Our heartfelt appreciation goes to Michael Onguech Costantino, MAMI Assistant, and Abraham Kuech Majok, Boma Health Worker and translator, for their invaluable contributions throughout the interview process. Special thanks are extended to all South Sudan Ministry of Health and MIHR collaborators in Juba and at the Gurei, Hai Dinka, Pariak, Wau, and Yambio primary healthcare centers.

## Authors contributions

HD and MHe designed the study, supported the interviews, managed and analyzed the data, and wrote the report. KD participated in the study design and reporting. ANB conducted the interviews.

## Patient and Public Involvement

Patients or the public WERE NOT involved in the design, or conduct, or reporting, or dissemination plans of our research.

